# Multilevel predictors categorization for post-CABG atrial fibrillation prediction

**DOI:** 10.1101/2025.07.06.25330976

**Authors:** Karina I. Shakhgeldyan, Vladislav Y. Rublev, Nikita S. Kuksin, Boris I. Geltser, Regina L. Pak

## Abstract

**Background:** Postoperative atrial fibrillation (PoAF) is known as common coronary artery bypass grafting (CABG) complication. Despite its association with increased risk of ischemic stroke, bleeding, acute renal failure and mortality there is still no ideal predictive tool with proper clinical interpretability.

**Methods:** A retrospective single-center cohort study enrolled 1305 electronic medical records of patients with elective isolated CABG. PoAF was identified in 280 (21.5%) patients. Prognostic models with continuous variables were developed utilizing multivariate logistic regression (MLR), random forest and eXtreme gradient boosting methods. Predictors were dichotomized via grid search for optimal cut-off points, centroid calculation, and Shapley additive explanation (SHAP). For multilevel categorization, we proposed to use threshold values combination identified during dichotomization, as well as ranking cut-off thresholds by MLR weighting coefficients (multimetric categorization method).

**Results:** Based on multistage selection, nine PoAF predictors were identified and validated. After categorization, prognostic models with continuous and multilevel categorical variables were developed. Multilevel categorical models advantage lies in their ability to explain PoAF prediction results and provide clinical interpretation, with comparable quality (AUC: 0.802 and 0.795).

**Conclusions:** Multilevel predictor categorization shows promise for PoAF predictions explanation, with the developed models demonstrating high accuracy and transparency in their conclusions.

**Highlights:**

1. Validation of new 1st diagnosed atrial fibrillation predictors were performed in patients with coronary heart disease after coronary artery bypass grafting with subsequent development of predictive models utilizing machine learning methods.
2. A new multilevel categorization method was tested, allowing to identify threshold values of predictors with the greatest predictive value, which were classified as risk factors for postoperative atrial fibrillation.
3. The best quality metrics (AUC - 0.802) were demonstrated by a stochastic gradient boosting prognostic model based on predictors identified by the multilevel categorization method.

## 1. Introduction

Postoperative atrial fibrillation (PoAF) affects 20-40% of patients after coronary artery bypass grafting (CABG) [1], with stable rates despite preventive strategies [2,3] while some authors even shows a potential trend to increase [4]. PoAF multiply the risk of stroke, bleeding, and renal failure fourfold, and doubles mortality at 30 days and 6 months [5]. The lack of a unified pathophysiological model has driven the creation of forecasting tools to personalize risk assessment [7–9].

Among PoAF prediction studies, the PoAF score [7] developed using MLR methods achieved an accuracy by an area under the ROC curve (AUC) of 0.63-0.65, with 0.6 sensitivity and 0.65 specificity values [8, 10]. Such limited accuracy prompted the use of new machine learning (ML) methods, allowing to improve model quality measured by AUC up to 0.7-0.75 [8, 11]. These models employed continuous and dichotomous predictors, with binary variables used to assess concomitant diseases. However, previous works lacked clinical justification for threshold values used in PoAF risk prediction. Multilevel categorization was only applied to age in some studies, with cut-off points set arbitrarily [7, 11].

Study aims to develop new prognostic models of PoAF in patients with coronary artery disease after isolated CABG based on preoperative predictors set and their multilevel categorization efficiency evaluation to improve prognosis quality and its clinical interpretation.

## 2. Material and methods

### 2.1 Data

Single-center cohort retrospective study results are presented, during which health records medical data of patients with coronary artery disease admitted for planned isolated CABG to the Vladivostok “Primorye Regional Clinical Hospital No. 1” cardiac surgery department from 2008 to 2023 were analyzed. Exclusion criteria was presence of atrial fibrillation (of any form) in anamnesis, as well as combination of CABG with any other surgery. Thus, the final dataset was represented by 1305 patients (992 men and 313 women) aged 35 to 83 years. The study protocol met local institutional requirements and received full approval; patient consent was not required. Far eastern federal university review board “Ethics approval: IRB protocol number: №1/3”, were approved on 19/03/2023. As the study involved a retrospective review of medical records, the requirement for patient consent was waived. The data was accessed from 19/03/23 to 12/08/23 for research purposes. During this period authors had access to information that could identify individual participants during data collection (DOB and medical record number).

First diagnosed PoAF episode was considered as the endpoint. AF episodes lasting more than 30 seconds, verified by the results of continuous electrocardiogram monitoring for at least 96 hours after CABG, were considered as PoAF development evidence. The PoAF presence was coded “1”, the absence – “0”. Thus, two patient groups were identified among the examined cohort. The first included 280 (21.5%) patients with AF paroxysms recorded during postoperative period in the hospital, the second - 1025 (78.5%) patients without cardiac arrhythmias.

The preoperative clinical and functional status of patients was assessed on the first day of hospital treatment by 130 factors, the main ones of which are presented in Appendix A. In addition to demographic, anthropometric, anamnestic data and physical examination results, clinical blood test indicators were analyzed. The diameters of the left (LAD) and right (RAD) atria, longitudinal dimensions of the left (LAL) and right (RAL) atrium, end-systolic (ESD) and diastolic (EDD) dimensions of the left ventricular (LV), ejection fraction (LVEF), and mean pulmonary artery pressure (MPAP) were determined. The ECG results were also analyzed: duration of P wave and QRS complex, PQ, QT intervals and RR.

### 2.2 Statistical Methods

Continuous characteristics distribution according to the Kolmogorov-Smirnov test differed from normal, so consequently nonparametric mathematical statistics methods were used for them. The indicators were presented as median (Me) and interquartile ranges (Q1; Q3), the Mann-Whitney test was used for continuous variables intergroup comparisons, and χ2 for categorical ones. For binary variables, odds ratios (OR) and their 95% confidence interval (CI) were calculated by Fisher’s exact test. Differences were considered statistically significant at p-value<0.05.

### 2.3 Machine Learning

PoAF predictive models were developed using MLR, random forest (RF) and eXtreme gradient boosting (XGB) methods. Their quality was assessed by 6 metrics: AUC, sensitivity, specificity, F1-score, positive predictive value (PPV) and negative predictive value (NPV). For optimal hyperparameters selection, the Grid Search Cross-Validation (GridSearchCV) optimization method from sklearn Python library was used.

The dataset was splitted into 2 samples: for training and cross-validation (80%) and for final testing (20%). The training and cross-validation procedure was performed by stratified k-Fold technique on 10 folds. The average AUC quality metric was used for best model selection, predictors picking and validation, and optimal hyperparameters selection by searching through a grid of acceptable values (GridSearchCV). For final testing, the best MLR, RF and XGB models with optimal parameters and hyperparameters were trained on 80% of the dataset, and tested on the final testing sample (20%). For quality metrics confidence, the assessment procedure was repeated 500 times, followed by metrics averaging, performing the initial division randomly using the bootstrapping method (Figure 1). Models were developed by utilizing open-source libraries Python version 3.9.16 (scikit-learn version 0.24.2, xgboost version 1.5.1).

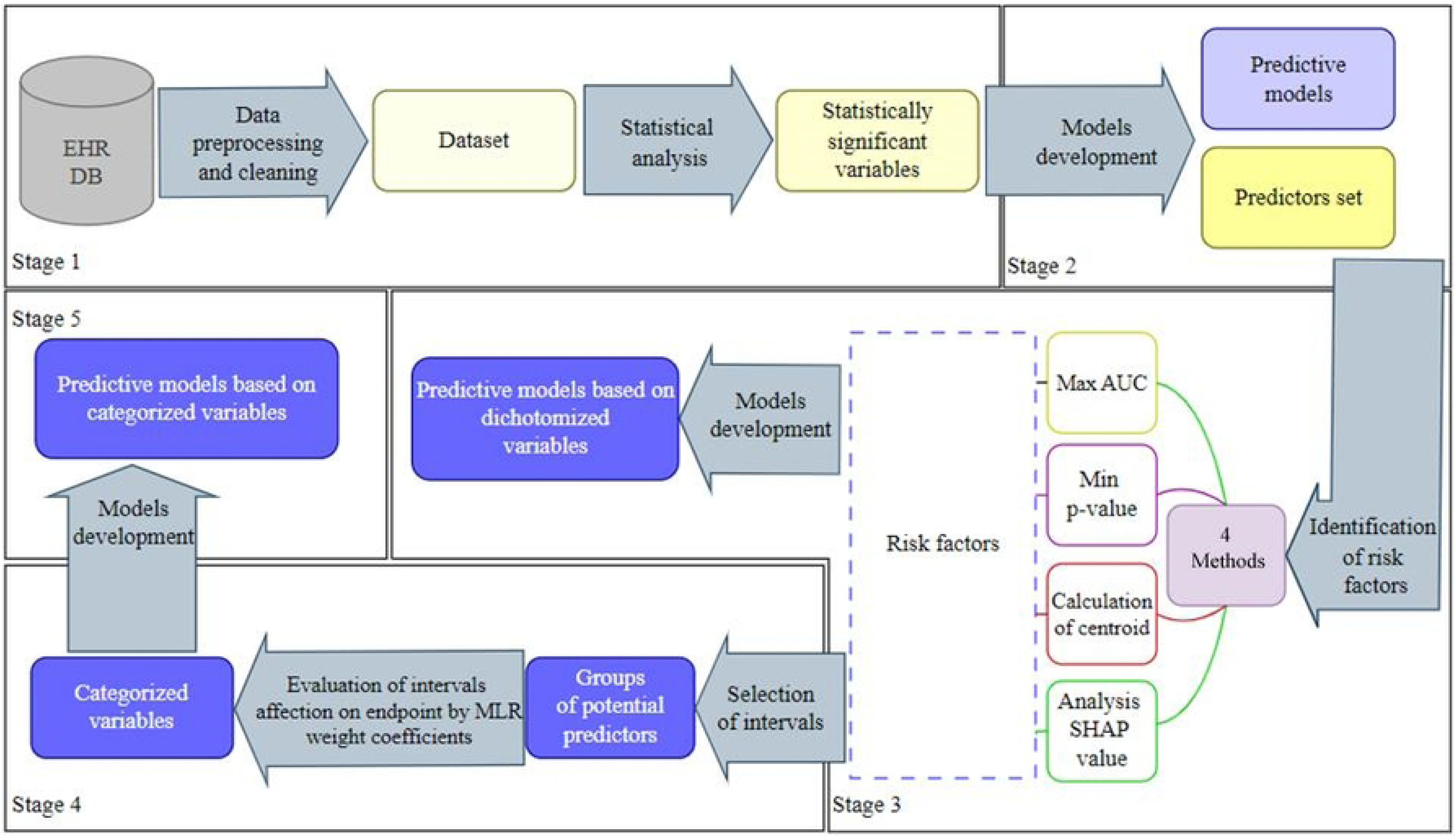

### 2.4 Variables categorization

This study utilized a multilevel categorization method that was previously reported by the authors [12].

To dichotomize potential predictors, we used grid-step optimization methods **Δ**=(max-min)/100: p-value minimization - Min(p-value), AUC maximization - Max(AUC), quartile method [13], centroid method and SHapley Additive exPlanation (SHAP) [14]. The Shapley method allowed us to identify thresholds at which the predictor influence function on the endpoint demonstrated singularity, which can be observed several times during the range of changes in the continuous attribute values [12]. To carry out multi-level categorization, we combined all threshold values identified with indicators dichotomization utilizing various methods, including the SHAP method. In this case, close threshold values were combined into one by averaging. The centroid method assumed usage of the analyzed characteristics median in the comparison groups (with and without PoAF) and values equidistant from them (centroids), with the help of which 4 categories were identified for each indicator [14]. The quartile method involves identifying 4 categories for each variable based on the results of assessing their medians, 2nd and 3rd quartiles [15].

For indicators endpoint influence degree assessment, the SHapley Additive exPlanation method was used.

### 2.5 Study design

The study design included 5 stages. At the very first of them utilizing intergroup comparisons tests, a potential PoAF predictors pool was formed. At the second stage of the study, PoAF prognostic models with predictors in a continuous form were developed by ML methods. The prognostic significance of the predictor was confirmed by AUC value increase after its inclusion in the model. During models development, all variables were considered, regardless of statistically significant differences in comparison groups, and hyperparameters were adjusted at the same stage. Models development and cross-validation was carried out on 80% of the dataset (derivation cohort), and the final testing was carried out on 20% (validation cohort). For further steps, the predictors and hyperparameters obtained at this stage were utilized. At the third stage, using various threshold values identification methods, binarization of continuous variables was carried out using a derivation cohort, and on their basis, PoAF prognostic models were developed, which were validated on the validation cohort. At the fourth stage of the study, multi-level categorization of variables was carried out using 4 approaches. In the first of them, only thresholds identified by the SHAP method were taken into account; in the second, the set of threshold values obtained by other dichotomization methods was expanded. In addition, thresholds obtained by the centroid method were considered, taking into account the medians of the groups with and without PoAF, as well as using quartiles Q1, Q2 and Q3. For risk factors endpoint influence degree assessment, MLR models were developed, the weights of which were used to code multilevel categorical predictors. Risk factors with negative or close to 0 weight coefficients in the MLR model were excluded from consideration. At the fifth stage of the study, 4 new PoAF prognostic models were developed by XGB method, the predictors of which were obtained by different methods of multilevel categorization. To assess statistically significant differences in quality metrics obtained by bootstrapping (n=500), 95% CI and Mann-Whitney test comparison results were used.

## 3. Results

### 3.1 Subject Characteristics

Intergroup analysis of clinical, demographic and laboratory parameters demonstrated that patients with PoAF were distinguished by older age, an increased prevalence of tricuspid regurgitation (TR) among them, lower levels of platelets, total protein and triglycerides in the blood. Individuals in this group had higher values of LV ESD, LAD, RAD and RAL, Ao/LV systolic pressure gradient and an increased duration of the QT and PQ intervals (Appendix A).

### 3.2 Machine Learning Models

During second stage, PoAF prognostic models were developed, validated and tested utilizing RF, XGB and MLR methods. For all models, the best AUC metric results were obtained by usage of ECG indicators (duration of QRS, QT, PQ, RR and P wave intervals), age, RAD, ESD, and TR as predictors. Developed models predictive value comparison showed that the XGB and RF methods provide higher forecast accuracy compared with MLR (AUC - 0.795 and 0.779 vs 0.698) (Table 1). Appendix B shows MLR model weight coefficients.

**Table 1.** Assessment of the accuracy of prognostic models for PoAF using predictors in continuous form.

| Metrics | Cross-validation |  |  | Final testing |  |  |
| --- | --- | --- | --- | --- | --- | --- |
|  | MLR | XGB | RF | MLR | XGB | RF |
| AUC | 0.698[0.697;0.699] | 0.774 [0.773; 0.775] | 0.77 [0.768; 0.771] | 0.698 [0.695; 0.702] | 0.795[0.791; 0.798] | 0.779[0.775; 0.782] |
| Sen | 0.643[0.641;0.644] | 0.706 [0.703; 0.708] | 0.689 [0.687; 0.691] | 0.643[0.636; 0.65] | 0.718[0.711; 0.725] | 0.7[0.694; 0.707] |
| Spec | 0.65[0.649;0.652] | 0.716 [0.714;0.717] | 0.695 [0.694; 0.697] | 0.65[0.647;0.654] | 0.72[0.716; 0.723] | 0.7[0.697; 0.704] |
| PPV | 0.31[0.309;0.311] | 0.384[0.383;0.386] | 0.363[0.361; 0.364] | 0.308[0.305; 0.311] | 0.394[0.391; 0.397] | 0.371[0.369 0.375] |
| NPV | 0.883[0.883;0.884] | 0.908[0.908;0.909] | 0.901[0.901; 0.902] | 0.884[0.882; 0.885] | 0.911[0.909; 0.913] | 0.903[0.901; 0.905] |
| F1-score | 0.416[0.415;0.418] | 0.495[0.493;0.497] | 0.473[0.471; 0.474] | 0.416[0.412; 0.42] | 0.507[0.503; 0.511] | 0.485[0.481; 0.488] |

### 3.3 Categorization

During third stage, PoAF predictors in a continuous form were dichotomized utilizing searching for the optimal cutoff threshold on the grid methods (Min(p-value) and Max(AUC)), along with the SHAP and centroid calculation method (Table 2). Threshold values usage, deviation from which is associated with PoAF likelihood increase, allows us to consider binarized data as risk factors for adverse events. The risk factor is coded “1” if the predictor value exceeds the threshold with the postfix “+” or does not reach it - with the postfix “-”, in other cases - “0”.

**Table 2.** PoAF continuous predictors dichotomization using different methods.

| Predictors | Min(p-value) | Max(AUC) | Centroid | SHAP |
| --- | --- | --- | --- | --- |
| Age, years | 60.0+ | 60.0+ | 64.0+ | 61+ |
| LV ESD, cm | 3.0+ | 3.0+ | 3.35+ | [3.1; 4.1]<br>5+ |
| RAD, cm | 4.12+ | 4.12+ | 4.4+ | [4.2; 5.3] |
| QRS, ms | 89- | 89- | 80- | 80- |
| QT, ms | 420+ | 382+ | 390+ | 390+ |
| PQ, ms | 163+ | 163.0+ | 155.0+ | [170;210] |
| RR, ms | 882.0+ | 882.0+ | 925.0+ | [700; 750]<br>[880; 1000]<br>1100 |
| P, ms | 120+ | 100+ | 100+ | 130+ |

**Abbreviations:** LV - left ventricle; LV ESD - end systolic dimension, RAD - right atrium transverse size.

Study results showed that the threshold values obtained by different binarization methods sometimes differed from each other. For example, the cutoff point for QRS according to SHAP was 80 ms, while when maximizing AUC, the cutoff point was fixed at 89 ms, and for the P wave, values above 130 ms were risk factors, while for Max(AUC) - above 100 ms (Table 2). The first three dichotomization methods considered isolated indicators and did not take into account predictive models. The SHAP method were applied to a multifactorial XGB model and the threshold value were defined as the point where the shap-value exceeded the level of 0.2 arbitrary units Thus, due to dichotomization, the following PoAF risk factors were identified: age over 61 years, RAD more than 4.2 cm, ESD - 3.1 cm, QRS duration less than 80 ms, QT more than 390 ms, P - 130 ms, PQ - 170 ms, RR - 700 ms (Figure 2). Annotation: The blue and red dotted lines indicate the cutoff thresholds. Abbreviations: LV - left ventricle, ESD - LV end systolic dimension, RAD - right atrium transverse size.

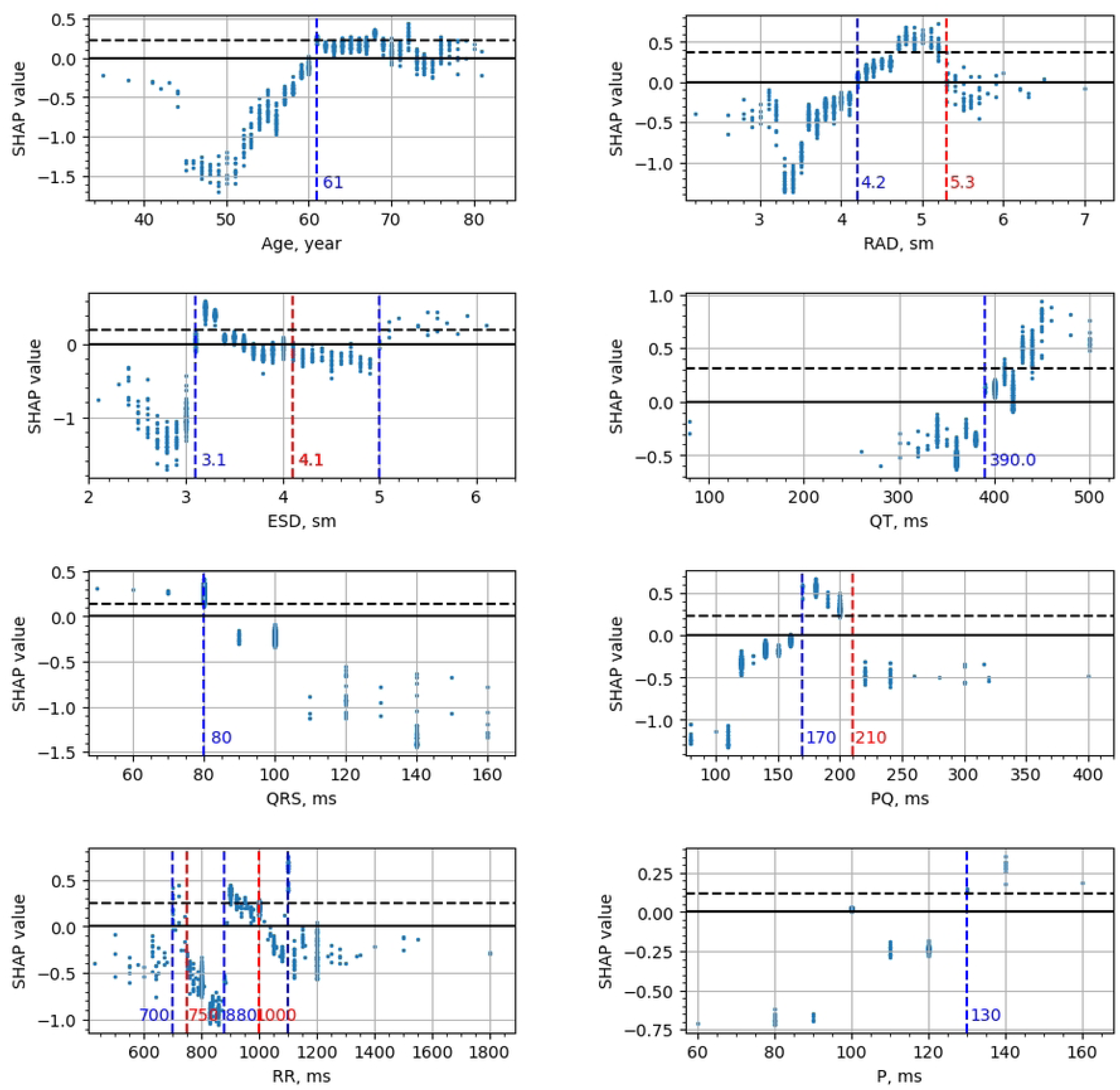

Using the QRS diagram as an example (Figure 2), it can be seen that the probability of PoAF developing in the range from 40 to 80 ms remains consistently high, but sharply decreases at its values ≥ 90 ms. Exceeding the QT parameter value more than 390 ms increases the risk of arrhythmia, but its maximum probability is fixed when the QT value is above 450 ms. Assessing the dynamics of changes in shap-value allows us to explain the relationship between various predictor values and study endpoint, which was the basis for utilizing this method in multilevel categorization procedures.

At the fourth stage of the study, utilizing various multilevel categorization methods, 4 groups of PoAF risk factors were formed. The first pool of risk factors was obtained from the shap-value analysis results (Figure 2). The second pool expanded the first due to threshold values obtained at the third stage of the study by several dichotomization methods. The third group of risk factors was represented by the predictors medians in the comparison groups and their centroids, and the fourth group used threshold values corresponding to predictors quartiles. To encode multilevel categorical predictors values, we used the weight coefficients (WC) of the MLR models developed for each risk factors group (Table 3). We call the approach that ensures the formation of a second pool of risk factors and their WC - multimetric categorization.

**Table 3.** Predictors weight coefficients and thresholds obtained by multilevel categorization methods.

| Predictors | SHAP |  | Multimetric categorization |  | Group medians and centroids |  | Quartiles |  |
| --- | --- | --- | --- | --- | --- | --- | --- | --- |
|  | Thresholds | WC | Thresholds | WC | Thresholds | WC | Thresholds | WC |
| Age, years | 61+ | 0.630 | [60; 64]<br>64+ | 0.621<br>0.689 | [63; 66]<br>66+ | 0.353<br>0.36 | [58; 64]<br>[64; 69]<br>69+ | 0.748<br>0.33<br>0.851 |
| ESD, cm | [3.1; 4.1]<br>5+ | 1.028<br>1.226 | [3.1; 4.1]<br>5+ | 1.0<br>1.15 | [3.3; 3.35]<br>3.35+ | 0.828<br>0.167 | [3.1; 3.3]<br>[3.3; 3.8]<br>3.8+ | 1.931<br>1.175<br>1.421 |
| RAD, cm | [4.2; 5.3] | 0.457 | [4.2; 5.3] | 0.533 | [4.3; 4.5]<br>4.5+ | 0.558<br>0.649 | [3.8.4.3]<br>[4.3; 4.8]<br>4.8+ | 0.906<br>1.377<br>0.877 |
| QRS, ms | 80- | 0.918 | 80- | 0.942 | 80- | 0.98 | [80; 100] | 1.302 |

MULTILEVEL AF PREDICTORS CATEGORIZATION
| Predictors | SHAP |  | Multimetric categorization |  | Group medians and centroids |  | Quartiles |  |
| --- | --- | --- | --- | --- | --- | --- | --- | --- |
|  | Thresholds | WC | Thresholds | WC | Thresholds | WC | Thresholds | WC |
| QT, ms | 390+ | 1.033 | 390+ | 1.027 | [380; 400]<br>400+ | 0.672<br>1.028 | [360; 395]<br>[395; 420]<br>420+ | 0.598<br>0.269<br>1.05 |
| RR, ms | [700; 750]<br>[880; 1000]<br>1100 | 1.306<br>1.211<br>2.327 | [700; 750]<br>[880; 1000]<br>1100 | 1.286<br>1.231<br>2.406 | [900; 950]<br>950+ | 1.256<br>0.629 | [800; 920]<br>[920; 1080]<br>1080+ | 0.324<br>0.398<br>0.429 |
| PQ, ms | [170; 210] | 0.991 | [170; 210] | 1 | 160+ | 0.523 | [140; 160]<br>[160; 180]<br>180+ | 0.658<br>1.712<br>0.864 |
| P, ms | 130+ | 1.547 | [100; 130]<br>130+ | 2.32<br>3.35 | 100+ | 5.886 | 100+ | 4.55 |
| TR | 1 | 0.683 | 1 | 0.703 | 1 | 0.785 | 1 | 0.721 |

**Abbreviations:** WC - weight coefficient; LV- left ventricular; ESD - LV end systolic dimension, RAD - right atrium transverse size, TR - Tricuspid regurgitation.

### 3.4 Models based on multilevel categorical predictors

At the fifth stage of the study, based on multilevel predictors obtained by various methods, 4 PoAF prognostic models were developed by XGB (Table 4). The best predictive properties were demonstrated by the model with predictors identified by the multilevel categorization method (AUC 0.802). The latter had comparable accuracy with the models including continuous variables or risk factors obtained by the SHAP method (AUC 0.802 vs. 0.795). A comparative accuracy assessment between prognostic models with predictors identified by dichotomization (Appendix C) and by multimetric multilevel categorization methods demonstrated the advantages of the latter, which was confirmed by statistically significant AUC metric differences (p-value <0.001). Besides, it allowed us to explain PoAF prognosis based on assessment results of predictors threshold values and WC (Table 3). Taking these data into account, it was concluded that in patients with coronary artery disease after CABG, the greatest likelihood of PoAF developing is associated with a P wave duration of ≥ 130 ms (WC - 3.35) and in the range [100-130 ms] with WC - 2.32, RR ≥ 1100 ms (WC - 2.41), as well as with RR in the range of 700-1100 ms, QT above 390 ms, PQ from 170 to 210 ms and QRS ≤ 80 ms. PoAF correlation was established for ESD in the range from 3.1 to 4.1 cm and above 5 cm, RAD - from 4.2 to 5.3 cm, age over 60 years and TR. The assessment of individual PoAF predictors’ influence on its development was performed utilizing the SHAP and XGB methods (Figure 3). The strongest influence is demonstrated by the QT indicator (shap-value 0.94), TR presence, the duration of the RR intervals and RAD. A low PoAF development probability is associated with the younger age (patients under 60 years), ESD below 3 cm and RAD below to 4.1 cm, PQ interval less than 150 ms and QRS above 100 ms.

**Table 4.**
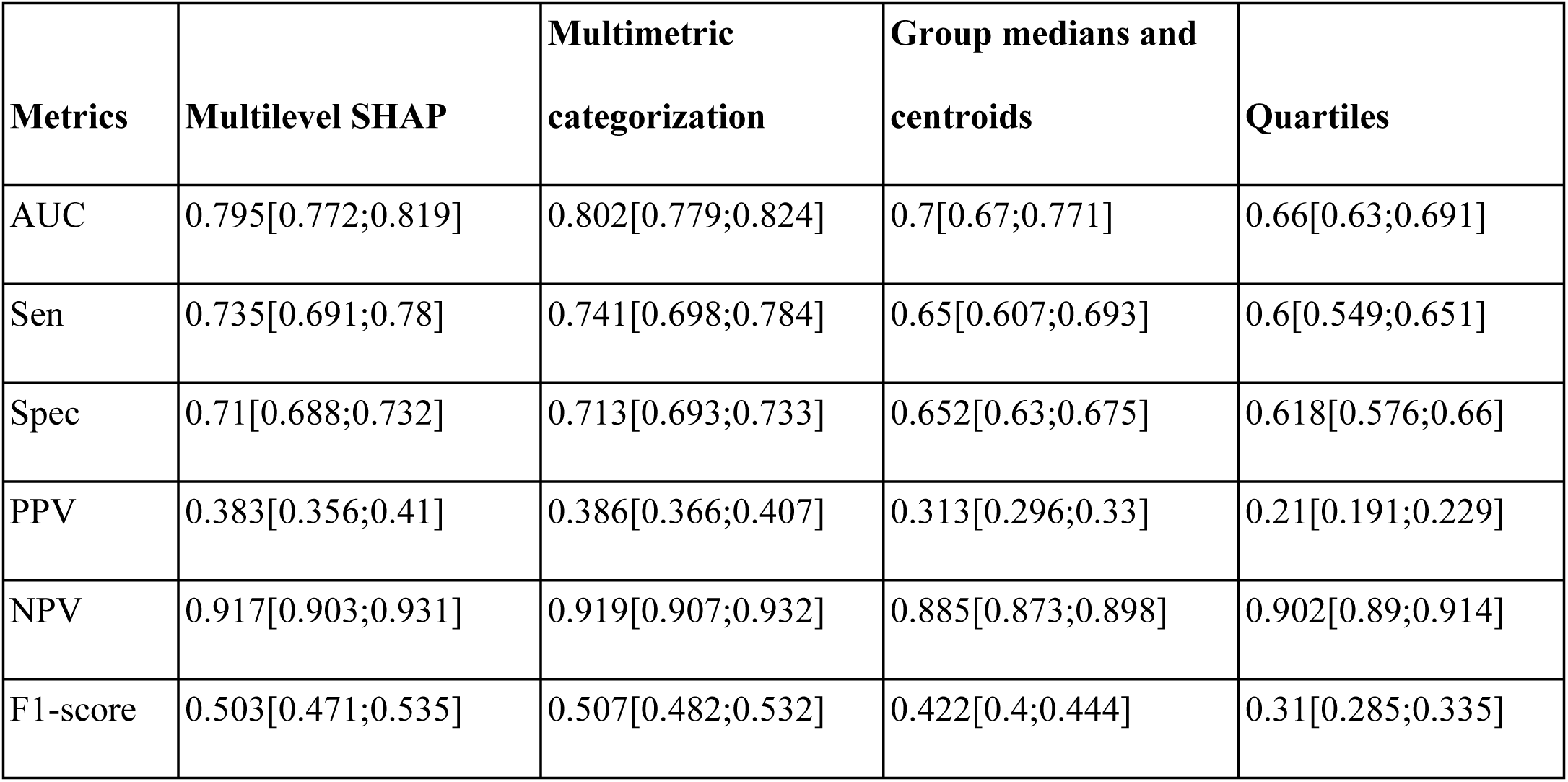
Accuracy assessment of PоAF prognostic models based on predictors with multilevel categorization.

| **Metrics** | **Multilevel SHAP** | **Multimetric categorization** | **Group medians and centroids** | **Quartiles** |
| --- | --- | --- | --- | --- |
| AUC | 0.795[0.772;0.819] | 0.802[0.779;0.824] | 0.7[0.67;0.771] | 0.66[0.63;0.691] |
| Sen | 0.735[0.691;0.78] | 0.741[0.698;0.784] | 0.65[0.607;0.693] | 0.6[0.549;0.651] |
| Spec | 0.71[0.688;0.732] | 0.713[0.693;0.733] | 0.652[0.63;0.675] | 0.618[0.576;0.66] |
| PPV | 0.383[0.356;0.41] | 0.386[0.366;0.407] | 0.313[0.296;0.33] | 0.21[0.191;0.229] |
| NPV | 0.917[0.903;0.931] | 0.919[0.907;0.932] | 0.885[0.873;0.898] | 0.902[0.89;0.914] |
| F1-score | 0.503[0.471;0.535] | 0.507[0.482;0.532] | 0.422[0.4;0.444] | 0.31[0.285;0.335] |

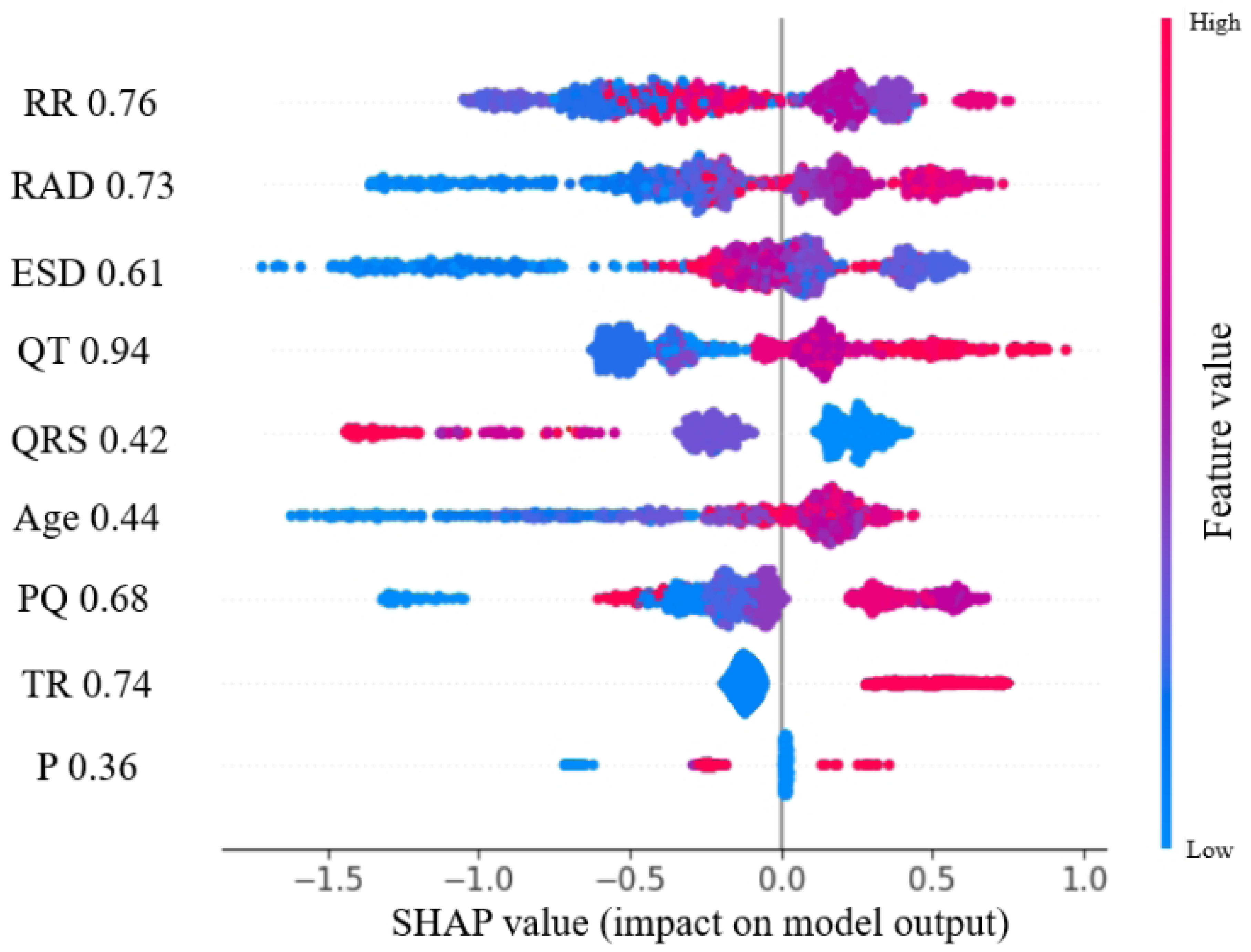

## 4. Conclusion

In recent years, prognostic models utilizing ML methods are being developed, wide usage of which in clinical practice is limited by the complexity of prognostic results interpretation. Promising tools for this problem solving are explainable artificial intelligence (XAI) algorithms, the elements of which includes the predictors threshold values determination and their ranking according to their influence intensity on the endpoint. Predictors threshold values determination is carried out using their categorization, which allows to detail the relationship between indicators of the clinical and functional patients status with the resulting variable. According to the literature, the most accessible method of multilevel categorization is descriptive statistics with the medians, quartiles or quantile calculation [16]. However, most of the categorization criticisms are associated precisely with this approach, which is primarily due to the dependence of such threshold values on a specific sample, lack of relationship with the clinical context, ignoring possible non-linear relationships, etc. [16]. An alternative method that takes into account the clinical context is searching for optimal threshold values based on minimization or maximization of objective functions, such as Min(p-value) or Max(AUC).

Recent literature about PoAF prediction problem analysis showed that utilizing categorization methods, PoAF risk factors were identified, which included the age of patients over 60 years [7, 17], 66 years [5, 18] or 70 years [19], increased LA size [5, 20], including with LAD > 4.5 cm [18] or > 3.9 cm [11] and reduced LVEF < 30% [7], increased P wave duration according to standard (>116 ms) [21, 22]. A number of anamnestic data are also validated as risk factors for PoAF: male gender [5, 23], the presence of arterial hypertension, chronic heart failure, chronic obstructive pulmonary disease, chronic kidney disease, diabetes mellitus [5], rheumatic heart disease [18, 23], mitral valve disease [3], previous cardiac surgery, metabolic syndrome and obesity [5, 6]. In our study, anamnestic features did not demonstrate predictive potential. The TR and RAD indicators were firstly verified as PoAF predictors. Our study confirmed the predictive value of age and ECG in relation to PoAF, but did not reveal a relationship between laboratory data and LVEF with the PoAF development.

Utilizing the patients with coronary artery disease after the CABG database example, we analyzed the effectiveness of various predictors threshold values searching methods, deviations from which increased their predictive potential and allowed to attribute them as PoAF risk factors. It was identified that the SHAP method, which considered as one of the promising XAI technologies, is a useful categorization tool due to the effective determination of cut-off thresholds, in particular for multilevel categorization and predictors relationship analysis both in continuous and categorical forms with the study endpoint. At the same time, multilevel categorical predictors obtained by combining SHAP data with other dichotomization methods results have been shown to provide higher predictive accuracy. Potential risks of information loss during new categorization methods usage were overcome by detailing knowledge about the interconnection of individual risk factors with the study endpoint. This was confirmed by predictive models quality criteria comparison for predictors both in continuous and multilevel categorical forms. Thus, for the best model with continuous predictors, AUC was 0.795, while utilizing multimetric categorization, it was 0.802.

## Dataset limitations

Study limitations are related to its retrospective nature, data usage from one medical institution, without model validation on patients from other hospitals. The dataset can be found at https://github.com/NikitaKuksin/DataSet_MultilevelPredictorsCategorizationPostCABG_AtrialFibrillation

## Data Availability

The dataset can be found at https://github.com/NikitaKuksin/DataSet_MultilevelPredictorsCategorizationPostCABG_AtrialFibrillation

## Supplemental Material

**Appendix A.** Clinical and functional characteristics of patients with coronary artery disease.

| Predictor | Group PoAF (n=280) |  | Group without PoAF (n=1025) |  | OR [95% CI] | p-value |
| --- | --- | --- | --- | --- | --- | --- |
| | Me (Q1; Q3)/abs | Mean ± SD/% | Me (Q1; Q3) | Mean ± SD | | |
| Age, years | 66 (61; 71) | 65.7 ± 6.9 | 63 (58; 69) | 62.9 ± 7.78 | - | <0.000001 |
| Female, abs. (%) | 76 | 27.14% | 237 (23.12%) |  | 1.24 [0.917; 1.673] | 0.188 |
| Height, cm | 170 (165; 176) | 170.1 ± 8.2 | 170 (165; 175) | 170 ± 8 | - | 0.802 |
| Weight, kg | 80 (73; 90) | 82.5 ± 14 | 80 (73; 90) | 82.1 ± 13 | - | 0.86 |
| BMI, $kg/m^2$ | 27.97 (25.3; 31.2) | 28.56± 4.85 | 28.1 (25.1; 31.2) | 28.51±4.69 | - | 0.736 |
| LVEF, % | 60 (54; 63.5) | 57.6 ±8.5 | 60 (51; 64) | 57.4 ±9.5 | - | 0.9 |
| RLVMI, c. u. | 1.01 (0.86; 1.18) | 1.05 ±0.307 | 0.98 (0.84; 1.16) | 1.02 ±0.264 | - | 0.263 |
| RTI, c. u. | 0.408 (0.363; 0.455) | 0.415 ±0.083 | 0.417 (0.37; 0.458) | 0.42 ±0.088 | - | 0.442 |
| LV ESD, cm | 3.3 (3.2; 3.7) | 3.49 ±0.58 | 3.35 (3; 3.8) | 3.45 ±0.59 | - | 0.185 |
| LV EDD, cm | 5.1 (4.8; 5.425) | 5.15 ± 0.58 | 5.1 (4.7; 5.4) | 5.1 ± 0.56 | - | 0.284 |
| Systolic pressure gradient Ao/LV, mm Hg | 7 (5; 9) | 8.67 ± 7.34 | 6 (5; 8) | 7.95 ± 7.88 | - | 0.048 |
| MPAP, mm Hg | 25 (21; 30) | 27.8 ± 8.97 | 25 (22; 28) | 26.9 ± 7.77 | - | 0.225 |
| LAL, cm | 3.9 (3.6; 4.2) | 3.93±0.56 | 3.9 (3.6; 4.3) | 3.98 ± 0.6 | - | 0.339 |
| LAD, cm | 4.6 (4; 5.1) | 4.57±0.74 | 4.3 (3.8; 4.9) | 4.38±0.73 | - | 0.0002 |
| Indexed LA volume, ml/m2 | 33.3 (24.5; 43.1) | 33.69±17.9 | 30.2 (23.7; 39.3) | 32.21±16.4 | - | 0.125 |
| RAL, cm | 3.8 (3.5; 4.1) | 3.8±0.5 | 3.7 (3.4; 4) | 3.71±0.52 | - | 0.0023 |
| RAD, cm | 4.5 (4.1; 4.9) | 4.5±0.64 | 4.3 (3.8; 4.8) | 4.28±0.66 | - | 0.0000033 |
| P, ms | 100 (100; 100) | 102.44±8.5 | 100 (100; 100) | 101.99±8 | - | 0.69 |
| PQ, ms | 160 (140; 180) | 161.3±32.7 | 150 (140; 180) | 156.8±36.7 | - | 0.025 |
| QRS, ms | 80 (80; 100) | 88.97± 14.97 | 80 (80; 100) | 94.4± 19.29 | - | 0.00036 |
| RR, ms | 950 (895; 1090) | 961.8±166 | 900 (800; 1080) | 943.7±166.5 | - | 0.129 |

MULTILEVEL AF PREDICTORS CATEGORIZATION
| Predictor | Group PoAF (n=280) |  | Group without PoAF (n=1025) |  | OR [95% CI] | p-value |
| --- | --- | --- | --- | --- | --- | --- |
| QT, ms | 400 (380; 430) | 396.8±34.7 | 380 (360; 420) | 387.9±38.5 | - | 0.00143 |
| Creatinine, $\mu\text{mol/l}$ | 92.83 (79.1; 110) | 95.3±25.7 | 97 (83; 110) | 98.7±24.2 | - | 0.036 |
| GFR, ml/min | 77.9 (62.6; 95.8) | 83.5±54.2 | 77.2 (63.6; 95.8) | 80.9±26.1 | - | 0,841 |
| CHF III-IV FC, abs. (%) | 43 (15.35%) |  | 124 (12.1%) |  | 1.3 [0.91; 1.93] | 0.156 |
| History of MI, abs. (%) | 32 (18.5%) |  | 143 (20%) |  | 0.91 [0.6; 1.4] | 0.75 |
| Stable angina pectoris III-IV FC | 65 (34.6%) |  | 247 (37.6%) |  | 1.14 [0.81; 1.61] | 0.478 |
| Extracardiac arteriopathy | 96 (34.3%) |  | 342 (33.3%) |  | 1.04 [0.79; 1.38] | 0.776 |
| AH, abs. (%) | 165 (95.38%) |  | 657 (91.76%) |  | 1.7 [0.81; 3.7] | 0.188 |
| Aortic stenosis, abs. (%) | 6 (2.1%) |  | 20 (1.95%) |  | 1.24 [0.49; 3.15] | 0.62 |
| TR, abs. (%) | 51 (18.21%) |  | 112(10.93%) |  | 1.8 [1.26; 2.6] | 0.00154 |
| MR, abs. (%) | 88 (31.4%) |  | 295 (28.78%) |  | 1.13 [0.85; 1.51] | 0.415 |
| AR, abs. (%) | 20 (7.15%) |  | 66 (6.4%) |  | 1.12 [0.66; 1.88] | 0.683 |
| CKD, abs. (%) | 74 (26.43%) |  | 262 (25.56%) |  | 1.04 [0.77; 1.41] | 0.758 |
| COPD, abs. (%) | 23 (13.29%) |  | 82 (11.45%) |  | 1.2 [0.72; 1.95] | 0.59 |
| DM, abs. (%) | 45 (26.01%) |  | 171 (23.88%) |  | 1.1 [0.77; 1.64] | 0.63 |
| Previous stroke, abs. (%) | 12 (6.94%) |  | 46 (6.42%) |  | 1.1 [0.56; 2.1] | 0.94 |
| Hemoglobin, g/l | 142 (131; 152) | 141±16.6 | 143(131; 153) | 141±16.9 | - | 0.77 |
| Red blood cells, $10^{12}/\text{l}$ | 4.69 (4.31; 5.03) | 4.65±0.58 | 4.69 (4.30; 5.06) | 4.65±0.58 | - | 0.98 |
| Leukocytes, $10^9/\text{l}$ | 6.8 (5.7; 8) | 7± 2 | 6.9 (5.8; 8.37) | 7.2±2.2 | - | 0.175 |
| Lymphocytes, $10^9/\text{l}$ | 1.87 (1.44; 2.42) | 1.9±0.73 | 2 (1.53; 2.53) | 2.1±0.92 | - | 0.09 |

MULTILEVEL AF PREDICTORS CATEGORIZATION
| Predictor | Group PoAF (n=280) |  | Group without PoAF (n=1025) |  | OR [95% CI] | p-value |
| --- | --- | --- | --- | --- | --- | --- |
| Platelets, 10<sup>9</sup>/l | 228 (182; 266) | 225±57 | 232 (192; 273) | 237±65 | - | 0.0325 |
| Total cholesterol, mmol/l | 4.35 (3.66; 5.4) | 4.6±1.36 | 4.45 (3.7; 5.43) | 4.7±1.41 | - | 0.132 |
| Glucose, mmol/l | 5.63 (5.11; 6.22) | 6±1.7 | 5.67 (5.13; 6.52) | 6.2±1.8 | - | 0.134 |
| Total protein, g/l | 70.9 (66.3; 73.7) | 69.6±7.3 | 71.4 (67.9; 75.2) | 71.1±7.8 | - | 0.00396 |
| Total bilirubin, µmol/l | 16.9 (12.1; 23.825) | 19.3±10 | 16.3 (11.7; 22.96) | 18.8±10.2 | - | 0.358 |
| Triglycerides, mmol/l | 1.48 (1.15; 1.88) | 1.68±1.1 | 1.63 (1.2; 2.2) | 1.82±0.94 | - | 0.0022 |
| Urea, mmol/l | 6 (5; 7.13) | 6.37±1.94 | 6 (4.97; 7.35) | 6.47±2.29 | - | 0.98 |
| Thrombin time, s | 19.6 (16.6; 21.5) | 20±6.14 | 19.9 (17.1; 21.4) | 20.1±7.2 | - | 0.67 |
| PTI, % | 93.6 (86; 99.7) | 92±14.58 | 94 (86.75; 102) | 94.26±25.3 | - | 0.172 |
| INR | 1.05 (1; 1.13) | 1.12±0.55 | 1.03 (0.98; 1.1) | 1.06±0.124 | - | 0.082 |
| SBP, mm Hg | 130 (130; 150) | 137±21 | 130 (125; 140) | 135±20 | - | 0.0193 |
| DBP, mm Hg | 80 (75; 80) | 80±8 | 80 (70; 80) | 79±8.5 | - | 0.00775 |
| Heart rate, beats/min | 68 (62; 75) | 70±12 | 68 (62; 72) | 69±10 | - | 0.178 |

**Abbreviations:** CI - confidence interval, BMI - body mass index, LV - left ventricle, LVEF - LV ejection fraction, RLVMI - relative left ventricular myocardial mass index, RTI - relative thickness index of left ventricle posterior wall, LV ESD - end systolic dimension, LV EDD - end diastolic dimension, MPAP - mean pulmonary artery pressure, LAL - left atrium medial-lateral size, LAD - left atrium anterior-posterior size, RAL - right atrium longitudinal size, RAD - right atrium transverse size, GFR - glomerular filtration rate, CHF - Congestive Heart Failure, FC- functional class, AH - arterial hypertension, TR - Tricuspid regurgitation, MR - mitral regurgitation, AR - aortic regurgitation, CKD - Chronic kidney disease, COPD- chronic obstructive pulmonary disease, DM - diabetes mellitus, PTI - prothrombin time index, INR- international normalized ratio, SBP - systolic blood pressure, DBP - diastolic blood pressure.

**Appendix B.** Weighting coefficients in a multivariate logistic regression model without predictors standardization.

| **Predictors** | **Weight coefficients** |
| --- | --- |
| Age | 0.036538 |
| RAD | 0.476447 |
| TR | 0.643446 |
| QRS | -33.214139 |
| QT | 12.580338 |
| RR | 0.551974 |
| PQ | 1.767283 |
| P | 14.158973 |
| ESD | 0.140284 |
| Intercept | -10.54463818 |

**Abbreviations:** RAD - right atrium transverse size, TR - Tricuspid regurgitation

